# Risk-stratified lifestyle intervention to prevent type 2 diabetes

**DOI:** 10.1101/2021.01.26.21249582

**Authors:** Andreas Fritsche, Robert Wagner, Martin Heni, Kostantinos Kantartzis, Jürgen Machann, Fritz Schick, Rainer Lehmann, Andreas Peter, Corinna Dannecker, Louise Fritsche, Vera Valenta, Renate Schick, Peter Paul Nawroth, Stefan Kopf, Andreas FH Pfeiffer, Stefan Kabisch, Ulrike Dambeck, Michael Stumvoll, Matthias Blüher, Andreas L Birkenfeld, Peter Schwarz, Hans Hauner, Julia Clavel, Jochen Seißler, Andreas Lechner, Karsten Müssig, Katharina Weber, Michael Laxy, Stefan Bornstein, Annette Schürmann, Michael Roden, Martin Hrabe de Angelis, Norbert Stefan, Hans-Ulrich Häring

## Abstract

**Background:** Lifestyle intervention (LI) can successfully prevent type 2 diabetes, but response to LI strongly varies depending on risk subphenotypes. We tested if individuals with prediabetes and a high-risk phenotype benefit from an intensification of LI.

**Methods and findings:** We conducted a risk stratified multicenter randomized controlled intervention study over 12 months with additional 2 year follow up. In eight University Hospitals in Germany, 1105 individuals (female 59%, age 58±11 years, BMI 31.1±6.0 kg/m^2^ (mean±SD)) with impaired fasting glucose and/or impaired glucose tolerance were included between May 2012 and May 2016 in the study. Participants were stratified into 2 groups; a high- and low-risk phenotype, based on insulin secretion, insulin sensitivity and liver fat content. Low-risk individuals were randomly assigned to conventional LI or control (1:1), high-risk individuals to conventional or intensified LI (1:1), each over one year. Intensified LI included doubling of physical exercise and time of counselling. The primary endpoint was change in post-challenge glucose levels, assessed by frequently sampled oral glucose tolerance tests. Secondary endpoints included changes in liver fat content, assessed by magnetic resonance spectroscopy. A total of 908 (82%) participants completed the study after 12 months of LI. In high-risk individuals, the mean difference estimate between conventional and intensified LI in change in post-challenge glucose levels from baseline was −0.290 mmol/l [CI: −0.544;−0.036], p=0.025. Liver fat content was more reduced by intensified LI than by conventional LI (mean difference estimate: −1.34 percentage points [CI: −2.17;−0.50], p=0.002), and cardiovascular risk decreased stronger with intensified LI than with conventional LI (mean difference estimate −1.82 [CI: −3.13−0.50], p=0.007). In low-risk individuals, conventional LI was not superior to control in reducing postprandial glucose, liver fat or cardiovascular risk. During the total observation period of 3 years, high-risk participants with intensified LI had a higher probability to normalize glucose tolerance compared to conventional LI (p=0.003). The limitations of this study include a relative short duration of LI, a non-completer rate of 18% and an underrepresentation of low risk individuals.

**Conclusions:** In high-risk individuals with prediabetes it is possible to improve glycemic and cardiometabolic outcomes by intensification of the commonly recommended conventional LI. Our results show that individualized, risk-phenotype-based LI can be implemented for the prevention of diabetes.

**Registration:** NCT01947595

**Author summary:** *Why Was This Study Done?:*

- Clinical trials in individuals with prediabetes have shown that the onset of type 2 diabetes can be delayed or prevented with lifestyle intervention.
- Among individuals with prediabetes, there is a large variability in the response to lifestyle intervention.
- It is unknown whether an intensification of intervention is able to improve the beneficial response.

*What Did the Researchers Do and Find?:*

- The present multicenter, risk stratified randomized and controlled intervention trial in 1105 German individuals with prediabetes prospectively confirms the existence of a high-risk prediabetes phenotype
- The intensification of lifestyle intervention in high-risk individuals improves the glycemic outcome after 1 year of lifestyle intervention, and additionally results in a higher frequency of regression to normal glucose tolerance after 3 years of follow up.
- .Intensification of lifestyle intervention results in a larger reduction of liver fat content and stronger improves cardiometabolic outcomes in high-risk individuals.

*What Do These Findings Mean?:*

- Strategies for the prevention of type 2 diabetes should include risk stratification and individualised interventions.
- Our results highlight a dose-effect relationship for lifestyle intervention and suggest that “one size fits NOT all” in the field of diabetes prevention.
- It remains to be clarified whether low risk individuals benefit from lifestyle intervention, as there was a low number of individuals in this risk group in the current study.

## Introduction

Lifestyle modification is the principal procedure for type 2 diabetes prevention in individuals with prediabetes. During the last two decades, multiple studies have shown that lifestyle intervention (LI) is effective in preventing diabetes. Several prospective randomized studies (1-4) have demonstrated that diabetes risk can be reduced by modifying diet and physical exercise. Such approaches yield relative diabetes risk reductions between 15% and 70% within 1 to 6 years of follow-up (5). Recent meta-analyses reported mean risk ratios of 0.35 (6), 0.57 (7) and 0.61 (8) when comparing LI to usual care. This points to a robust benefit of LI for the prevention of type 2 diabetes, which is sustainable and extends beyond the duration of the intervention (4,9,10).

Nevertheless, there is a pressing need for making LI more efficient for diabetes prevention because a considerable proportion of participants in LI trials do not benefit from the intervention. They are often referred to as “non-responders” (11,12). For example, every fifth patient of the LI group in the Diabetes Prevention Programme (DPP) developed type 2 diabetes within four years (2). An alternative definition of non-response is the inability to regress from prediabetes to a normal glucose regulation during a LI program (11). In the DPP, only ∼40% of participants accomplished regression to normal glucose regulation (11), i.e. 60% were LI non-responders. Furthermore, there is the important question whether lifestyle intervention is necessary in all individuals with prediabetes (13). There are individuals with prediabetes who do not progress to diabetes during 11 years of follow up even without intervention (14). In such individuals with “intermediate hyperglycemia”, lifestyle intervention with the sole purpose to lower blood glucose might be of less importance. Both observations of non-response to LI and non-progression to diabetes highlight the need for risk stratified intervention strategies in individuals with prediabetes.

The fundamental question is which phenotypes determine the risk for diabetes and especially the response and non-response to LI. A recent *post hoc* analysis of the DPP showed that response varies based on diabetes risk (15), suggesting an adaption of LI on individual risk. In a retrospective analysis of the Tuebingen Lifestyle Intervention Program (TULIP) study, we identified a high-risk phenotype associated with higher probability of short-term (16) and long-term non-response (12) to LI. This phenotype represents beta cell dysfunction and/or insulin resistant non-alcoholic fatty liver disease (NAFLD), which is also associated with increased cardiometabolic risk (17). Similar phenotypes have been recently identified by cluster analysis in manifest type 2 diabetes patients (18). They were associated with a severe disease course and higher risk for diabetes-related complications.

Therefore, it is crucial to improve the efficiency and effectiveness of LI programs in order to overcome non-response in high-risk participants and to avoid overtreatment in low-risk participants. We therefore designed a prospective risk-stratified randomized controlled multi-center LI study to answer the following questions:

I. Can non-response in high-risk individuals be overcome by intensification of LI?
II. Is lifestyle intervention effective in low-risk individuals with prediabetes?

## Methods

### Study design

The prediabetes lifestyle intervention study (PLIS) (ClinicalTrials.gov Identifier: NCT01947595) is a stratified randomized multi-center trial involving eight study sites in university hospitals in Germany (Appendix Table 1). The primary hypothesis is that individuals with prediabetes who have high risk for a failure to restore normal glucose regulation with conventional LI will benefit from an intensification of the LI. Prediabetes was diagnosed from fasting and 2 hour post-challenge glucose (2hPG) levels after a standardized oral glucose tolerance test (OGTT), according to the criteria of the American Diabetes Association (19). HbA1c was not used as a definition for prediabetes. Screening procedures also involved measurement of liver fat content, insulin sensitivity and insulin secretion. Based on previously established cut-off levels (14), these variables were used for risk stratification. Low-risk participants (LR) were randomized to receive no lifestyle intervention (control group, LR-CTRL) or a conventional (LR-CONV) lifestyle intervention. Participants with high-risk (HR) were randomized to receive either a conventional lifestyle intervention (HR-CONV) or an intensive lifestyle intervention (HR-INT). Randomisation was performed using a computer-based block-randomisation at the center of Tübingen by a study supervisor. For this, a self-devised randomiser with a permuted block randomization with a block size of 30 was used. At each study site, the study personnel was blinded, except for the principal investigator and the personnel performing the actual lifestyle councelling. The primary outcome measure 2hPG was assessed by an OGTT after 12 months, an intermediate OGTT was performed after 6 months. Secondary outcome measures were liver fat content, insulin sensitivity and secretion and cardiovascular risk. Participants were enrolled between 2012 and 2016. The study protocol was approved by all local ethics committees of the participating institutions. This study has been reporting in line with the CONSORT guidelines and the completed checklist is in the Supplementary Appendix. The detailed study protocol is available <u>online.</u>

**Table 1.**
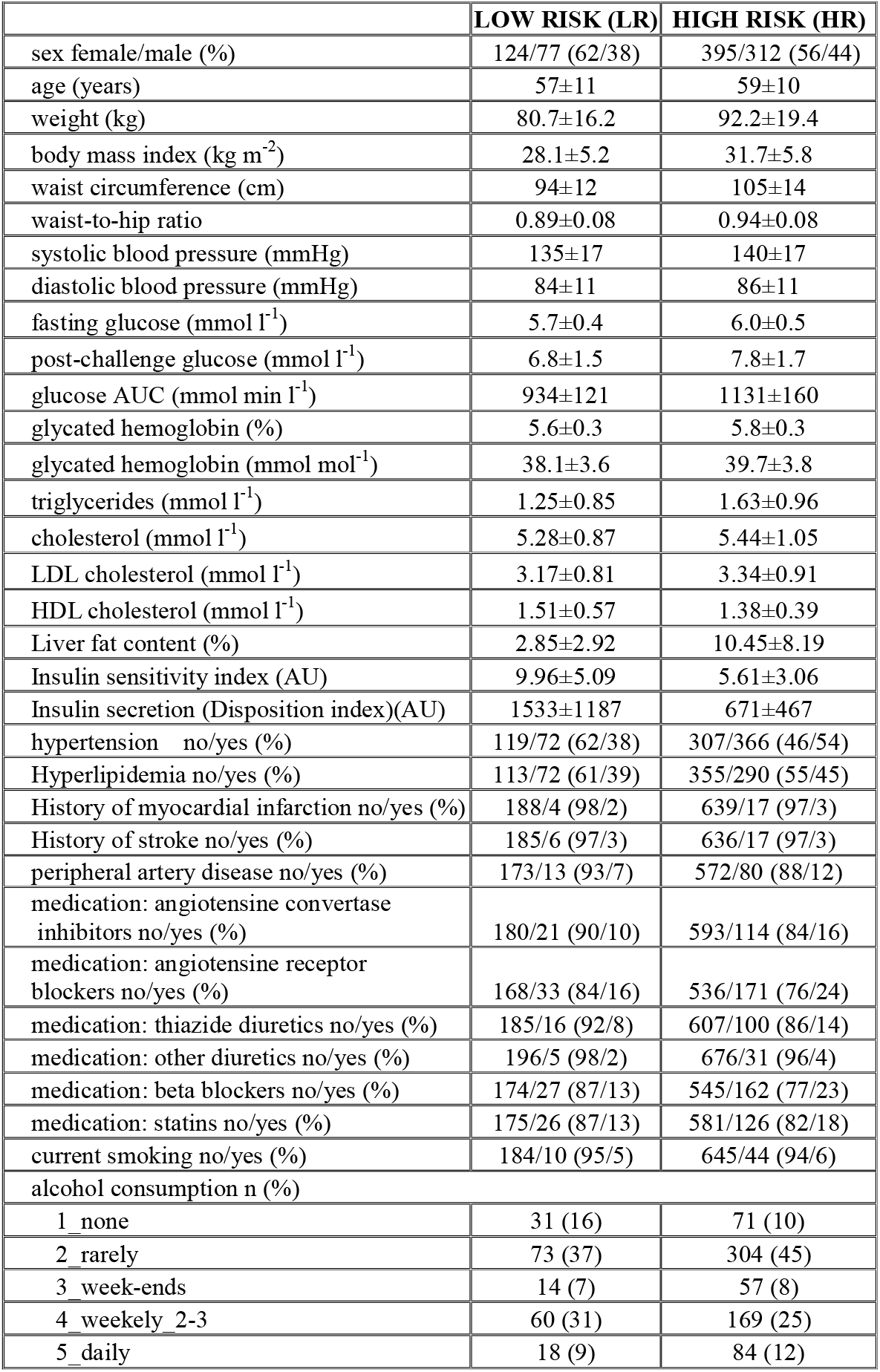
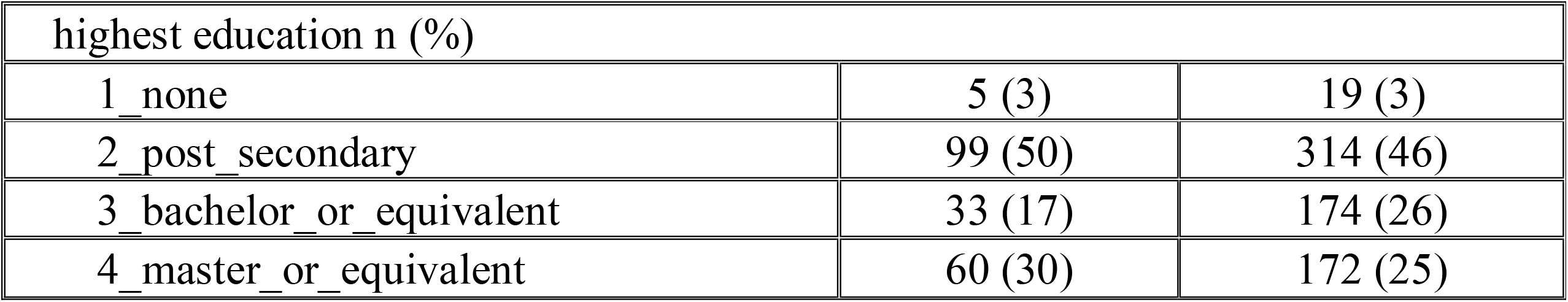
Comparison of baseline parameters (mean±SD) of the low-risk versus high risk group (individuals with complete follow-up).

|  | **LOW RISK (LR)** | **HIGH RISK (HR)** |
| --- | --- | --- |
| sex female/male (%) | 124/77 (62/38) | 395/312 (56/44) |
| age (years) | 57±11 | 59±10 |
| weight (kg) | 80.7±16.2 | 92.2±19.4 |
| body mass index (kg m<sup>-2</sup>) | 28.1±5.2 | 31.7±5.8 |
| waist circumference (cm) | 94±12 | 105±14 |
| waist-to-hip ratio | 0.89±0.08 | 0.94±0.08 |
| systolic blood pressure (mmHg) | 135±17 | 140±17 |
| diastolic blood pressure (mmHg) | 84±11 | 86±11 |
| fasting glucose (mmol l<sup>-1</sup>) | 5.7±0.4 | 6.0±0.5 |
| post-challenge glucose (mmol l<sup>-1</sup>) | 6.8±1.5 | 7.8±1.7 |
| glucose AUC (mmol min l<sup>-1</sup>) | 934±121 | 1131±160 |
| glycated hemoglobin (%) | 5.6±0.3 | 5.8±0.3 |
| glycated hemoglobin (mmol mol<sup>-1</sup>) | 38.1±3.6 | 39.7±3.8 |
| triglycerides (mmol l<sup>-1</sup>) | 1.25±0.85 | 1.63±0.96 |
| cholesterol (mmol l<sup>-1</sup>) | 5.28±0.87 | 5.44±1.05 |
| LDL cholesterol (mmol l<sup>-1</sup>) | 3.17±0.81 | 3.34±0.91 |
| HDL cholesterol (mmol l<sup>-1</sup>) | 1.51±0.57 | 1.38±0.39 |
| Liver fat content (%) | 2.85±2.92 | 10.45±8.19 |
| Insulin sensitivity index (AU) | 9.96±5.09 | 5.61±3.06 |
| Insulin secretion (Disposition index)(AU) | 1533±1187 | 671±467 |
| hypertension no/yes (%) | 119/72 (62/38) | 307/366 (46/54) |
| Hyperlipidemia no/yes (%) | 113/72 (61/39) | 355/290 (55/45) |
| History of myocardial infarction no/yes (%) | 188/4 (98/2) | 639/17 (97/3) |
| History of stroke no/yes (%) | 185/6 (97/3) | 636/17 (97/3) |
| peripheral artery disease no/yes (%) | 173/13 (93/7) | 572/80 (88/12) |
| medication: angiotensine convertase inhibitors no/yes (%) | 180/21 (90/10) | 593/114 (84/16) |
| medication: angiotensine receptor blockers no/yes (%) | 168/33 (84/16) | 536/171 (76/24) |
| medication: thiazide diuretics no/yes (%) | 185/16 (92/8) | 607/100 (86/14) |
| medication: other diuretics no/yes (%) | 196/5 (98/2) | 676/31 (96/4) |
| medication: beta blockers no/yes (%) | 174/27 (87/13) | 545/162 (77/23) |
| medication: statins no/yes (%) | 175/26 (87/13) | 581/126 (82/18) |
| current smoking no/yes (%) | 184/10 (95/5) | 645/44 (94/6) |
| alcohol consumption n (%) |  |  |
| 1_none | 31 (16) | 71 (10) |
| 2_rarely | 73 (37) | 304 (45) |
| 3_week-ends | 14 (7) | 57 (8) |
| 4_weekely_2-3 | 60 (31) | 169 (25) |
| 5_daily | 18 (9) | 84 (12) |

| highest education n (%) |  |  |
| --- | --- | --- |
| 1_none | 5 (3) | 19 (3) |
| 2_post_secondary | 99 (50) | 314 (46) |
| 3_bachelor_or_equivalent | 33 (17) | 174 (26) |
| 4_master_or_equivalent | 60 (30) | 172 (25) |

### Participants

Individuals participated in a screening OGTT if they had clinically suspected prediabetes, or at least 50 points in the German Diabetes Risk assessment battery (20). Basic inclusion criteria comprised age between 18 and 75 years and a BMI < 45 kg/m^2^ and diagnosis of impaired fasting glucose (IFG) and/or impaired glucose tolerance (IGT). Exclusion criteria are listed in Appendix Table 2. All participants provided written informed consent.

**Table 2:**
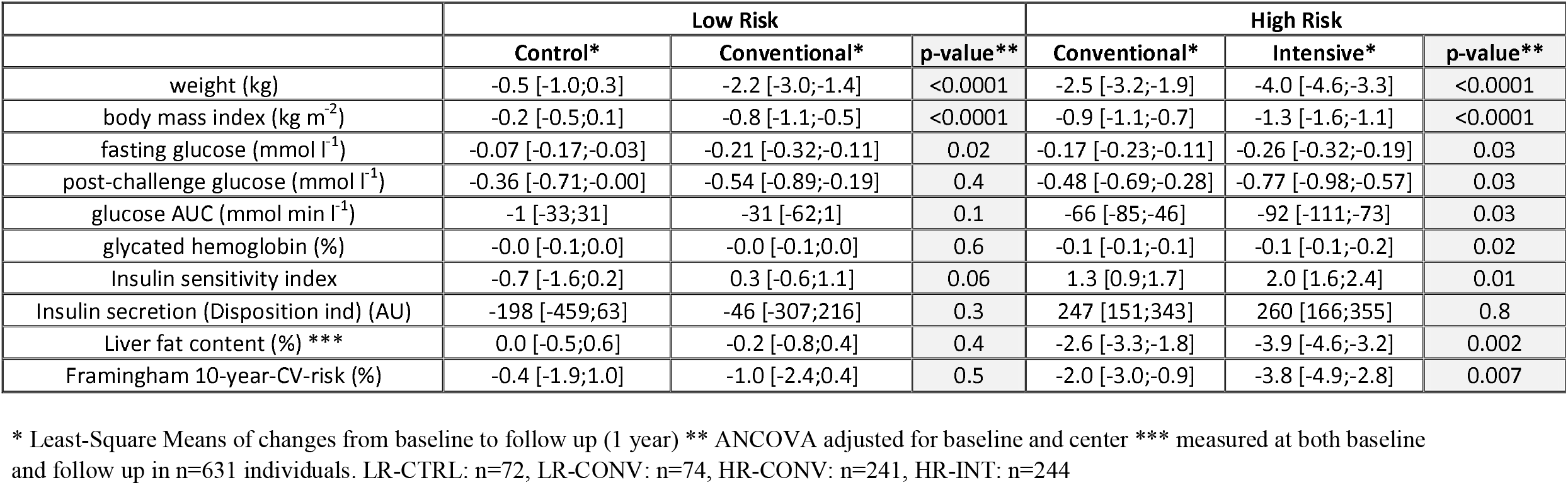
Changes of key study variables between baseline and follow up in low-risk (control vs conventional LI) and high-risk individuals (conventional vs intensive LI)

### Intervention

The duration of the LI was 12 months. In both the conventional and the intensified treatment groups, the lifestyle intervention was aimed at reaching a body weight reduction of 5% in participants with a body mass index (BMI) > 25 kg/m^2^ by reducing fat intake to less than 30% of total energy intake, reducing saturated fat intake to less than 10% of total energy intake, and increasing fiber intake to more than 15 g per 1000 kcal of total energy intake. Participants of the conventional intervention group received eight LI sessions in total over 1 year. They were advised to perform 3 hours of exercise weekly. Participants of the intensified LI group received 16 coaching sessions in total over 1 year with advice to exercise 6 hours weekly. The duration of the one-to-one coaching sessions was 30-60 minutes. They included dietary counselling based on diet protocols completed by the participants on four consecutive days. Furthermore, exercise counselling was performed on the basis of data from accelerometers, also enabling the assessment of accomplishing exercise goals. During each visit, lifestyle advisors graded adherence to intervention based on diet and exercise protocols. All dieticians/lifestyle advisors were trained using the same curriculum (10h) by a team from the primary site before stating recruitment. Refresher courses and face to face meetings between advisors were organized at least yearly between the study centers in workshops to ensure team building and harmonized counselling across all study sites involved.

Participants of the control group only received a single 30 minutes one-to-one consultation with a dietician at baseline.

### Oral glucose tolerance test and analytical procedures

OGTTs were performed at 8:00 after an overnight fast. Participants ingested 75 g glucose (Accu-Check Dextro O.G.T., Roche). Blood samples were obtained at fasting, 30, 60, 90 and 120 minutes via an indwelling venous catheter. Blood samples were immediately put on ice and frozen at −80°C.

Glucose levels were measured locally at the study sites. Plasma insulin was measured centrally in the laboratory of the Tübingen University Hospital with a commercial chemiluminescence assay on an ADVIA Centaur XP (Siemens Healthineers). Hepatic fat content was assessed by magnetic resonance spectroscopy or imaging (see below), whenever possible. For a small proportion of participants who were unable to undergo magnetic resonance studies or when magnetic resonance studies were not available, hepatic steatosis was assessed by ultrasound imaging to allow risk stratification if needed.

### Magnetic resonance imaging and spectroscopy

Liver fat content was determined by localized proton magnetic resonance spectroscopy (^1^H-MRS) using stimulated echo acquisition mode in the posterior hepatic segment 7 (21). Liver fat content was determined by the ratio of signal integrals of fat (methylen+methyl signal) and total signal (water+fat), expressed in %. ^1^H-MRS was not available in one center. Here, liver fat content was quantified by a chemical-shift selective imaging technique generating fat and water selective images (22). Liver fat content was determined from a manually drawn region of interest in segment 7, performed separately on the water selective and the fat selective image. Similar to the^1^H-MRS method, liver fat content was calculated as fat/(water+fat)∗100 correcting for relaxation effects in order to make the imaging approach comparable to MRS.

### Calculations and Statistical Analysis

The study was designed to be powered to detect a difference of 0.44 mmol/l in post-challenge glucose in a study population of 200 per intervention group. A complete cases approach was used for all statistical analyses. As a complementary analysis, we provide the main outcomes also after imputing all missing variables (see Appendix Table 3).

The primary and secondary endpoints were analyzed by general linear models. For example, as primary end-point, post-challenge glucose at the end of the intervention was evaluated with ANCOVA in dependence from intervention, adjusting for baseline post-challenge glucose and study center as fixed effect. Other tested variables were evaluated with similar models. For sensitivity analysis in the unbalanced high- and low-risk groups, we performed center-stratified, bias-corrected accelerated bootstrapping to estimate the confidence intervals of the intervention effect sizes on post-challenge glucose. Results from general linear models are provided as least-square means with 95% CI (Table 2). All other tables show means and standard deviation. Insulin sensitivity was calculated using glucose and insulin levels obtained during the OGTT with the equation of Matsuda and DeFronzo (ISI) (23). Insulin secretion was calculated with the insulinogenic index (IGI) (24). To obtain insulin secretion capacity adapted for actual insulin sensitivity, the disposition index (ISI×IGI) was used. In addition, we have conducted post-hoc tests using alternative insulin sensitivity and secretion variables to predict the primary outcome. The prediction power was very similar to our current approach, therefore we think that the kind of indices estimating insulin sensitivity and secretion do not critically influence our results. Cardiovascular risk was assessed with the Framingham risk score which was calculated using the equation provided by D’Agostino et al. (25), with participants having concomitant IFG and IGT being taken as participants with diabetes. All statistical analyses were performed in R (Version 3.4) (26).

## Results

### Study participants

Out of 2561 individuals with increased risk for diabetes, a total of 1160 individuals were identified as eligible, agreed to participate and underwent risk stratification into a low-risk group (LR) and a high-risk group (HR). A total of 1105 individuals were subsequently randomized into the four study groups and received allocated intervention. Details can be seen in figure 1.

**Figure 1.**
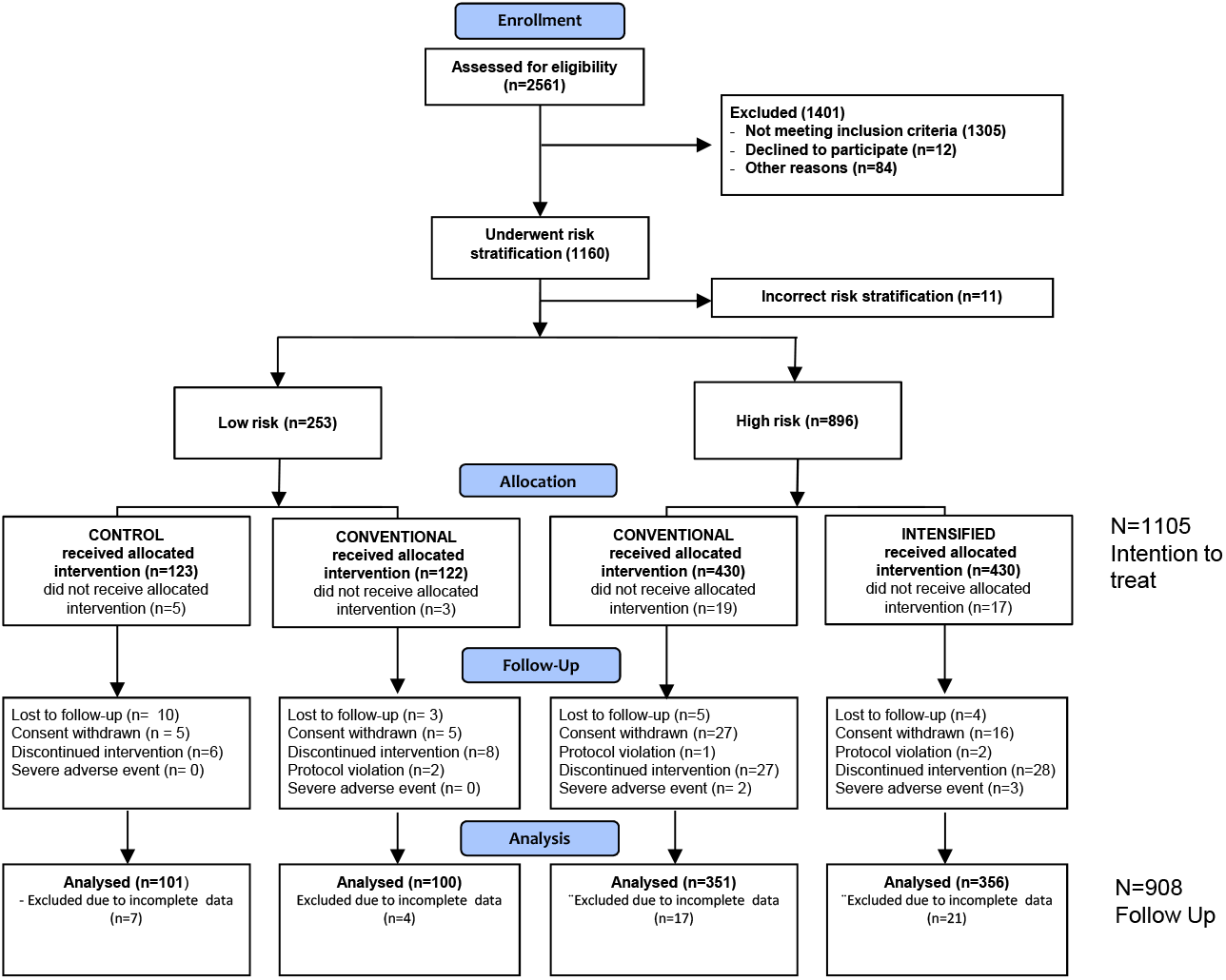
Participant flow during the study (Consort diagram).

After one year, 908 individuals (82%) completed the study, and outcome data for the primary endpoint (complete glucose data from OGTT) were obtained. Among these, high-risk subjects were significantly older and had higher BMI. They also differed in all major metabolic traits such as glucose and lipid levels, insulin sensitivity and insulin secretion (see Table 1). The randomization procedure resulted in balanced demographic and clinical characteristics between LR-CTRL and LR-CONV as well as between HR-CONV and HR-INT (see Appendix Table 4).

Non-completers did not differ from completers regarding the allocation to risk groups and intervention arms. Non-completers were significantly more often female, younger and had higher BMI (see Appendix Table 5).

### Primary outcome: post-challenge glucose

Post-challenge and fasting glucose levels decreased in all study groups (See Table 2).

In high-risk subjects, the mean difference estimate between conventional and intensified LI of the change of post-challenge glucose levels from baseline to 1 year follow-up was −0.290 mmol/l [CI: −0.544;−0.036], p=0.025, adjusted for baseline and center (see Figure 2). For the least-square means of changes from baseline to follow up in each intervention group, see Table 2. In low-risk subjects, the change in 2hPG was not significantly different between the LR-CTRL and LR-CONV groups (mean difference estimate, 0.188 mmol/l [CI: - 0.224;0.600], p=0.4, see Figure 2).

**Figure 2.**
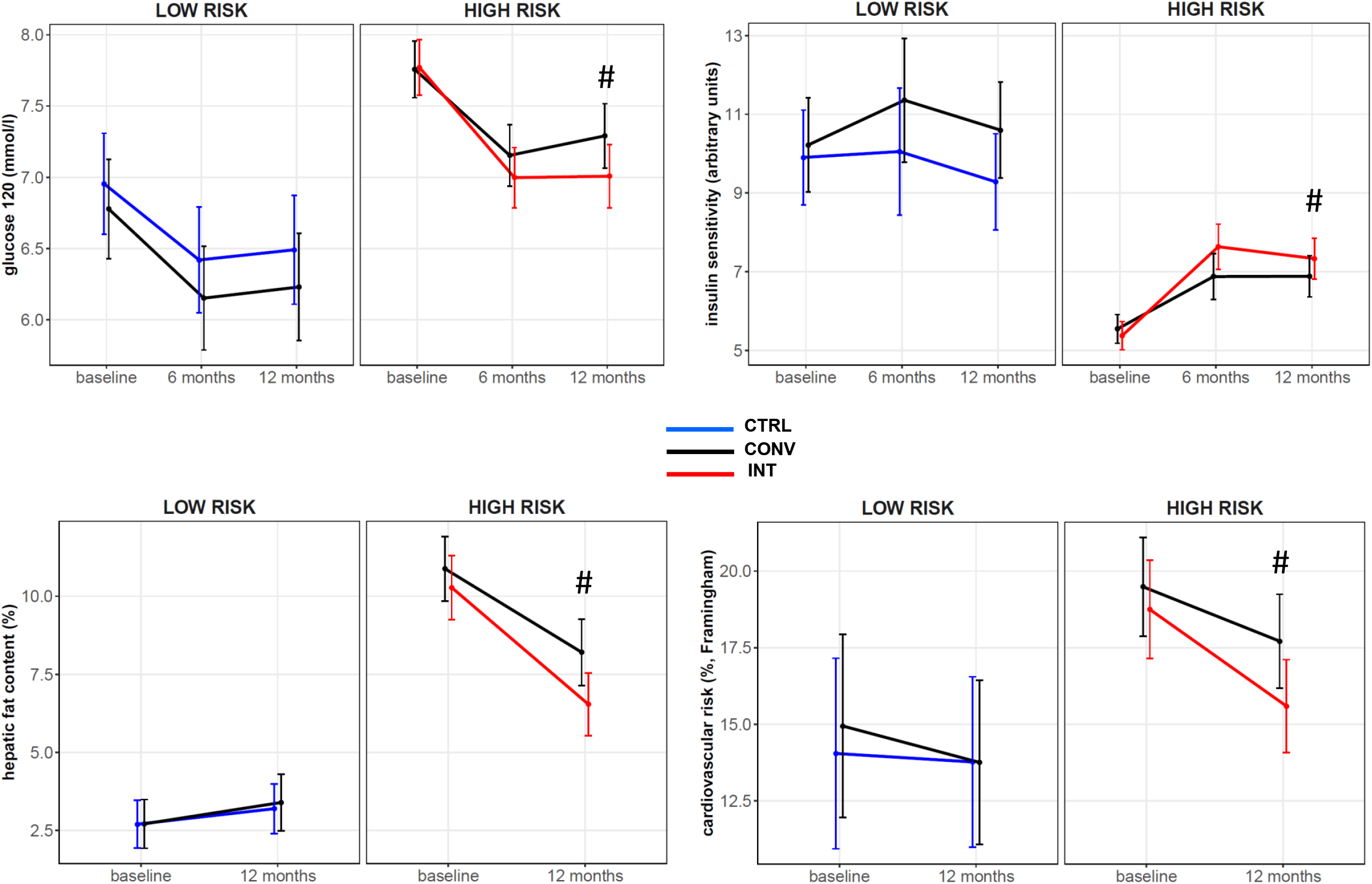
Plasma glucose levels at 120 minutes after standardized 75 g glucose challenge (panel A), and insulin sensitivity (panel B) at baseline, 6 and 12 months during LI, hepatic fat content (panel C), and cardiometabolic risk (panel D) at baseline and 12 months during LI. Values shown as least-square means (LSM) with 95% confidence intervals, adjusted for study center. # indicates significant difference (p<0.05) between HR-CONV and HR-INT in change of the parameter from baseline.

A sensitivity analysis using bootstrap showed a 2hPG-lowering effect estimate of −0.17 mmol/l [CI: −0.60; 0.19] for the conventional LI in the low-risk group, and an effect estimate of −0.30 mmol/l [CI: −0.57; −0.05] for the intensified LI in the high-risk group.

### Change in glycemic categories

We defined response to LI as improvement of glycemic category (see Appendix Figure 1). The rate of responders in the high-risk groups was higher in HR-INT compared to HR-CONV (53 vs 41%, p=0.001, rate ratio 1.29 [CI: 1.08;1.51]). High-risk individuals with intensive LI had a 63% increased probability to respond to LI compared to their conventionally treated counterparts (OR 1.63, 95% CI 1.21;2.19). No significant difference was found in participants with low risk (see Appendix Figure 1). The flow between glycemic categories during the study is visualized in Appendix Figure 2.

### Regression to normal glucose tolerance during long-term follow up

Subsequent to the LI, follow-up visits with OGTT were performed 1 and 2 years later. During this total observation period of 3 years, intensive LI lead to a cumulative higher conversion rate to normal glucose tolerance in high risk individuals compared to conventional LI (p=0.003, parametric proportional odds survival model using an exponential baseline risk distribution, Figure 3). In low-risk individuals, participants receiving conventional lifestyle had a higher chance of conversion to normal glucose tolerance compared to controls during 3 years of follow-up (p=0.01).

**Figure 3.**
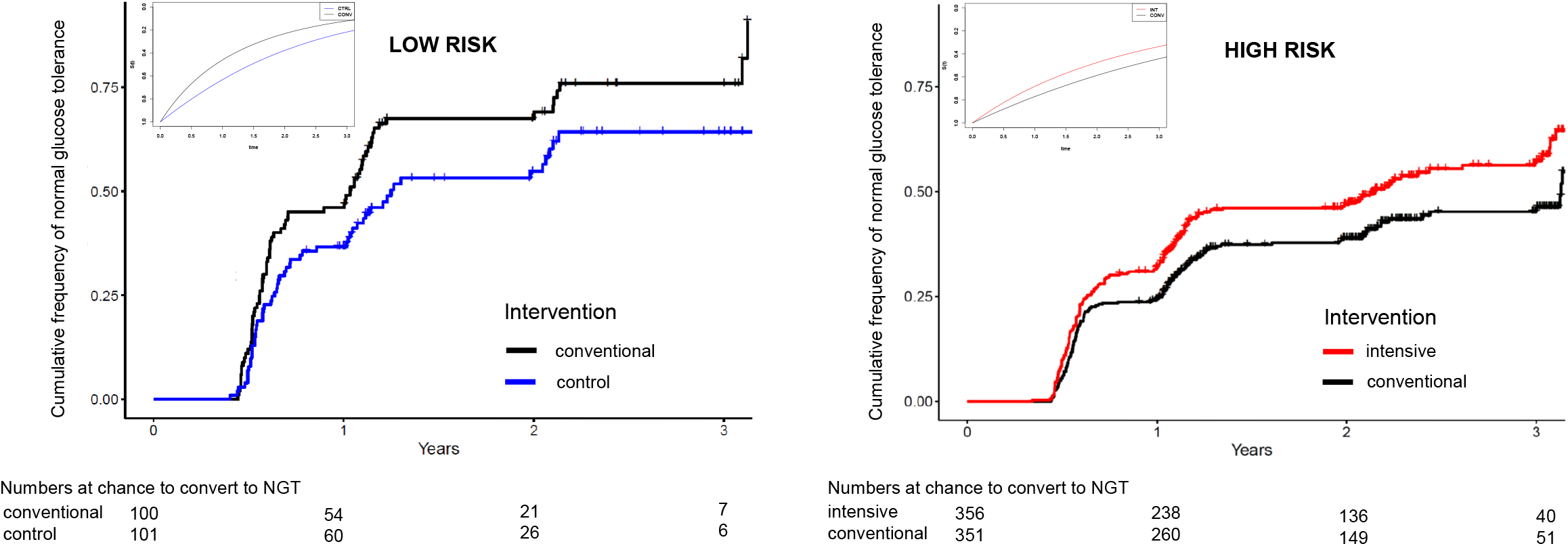
Results after 3 years observation (1 year of lifestyle intervention) and additional 2 years of follow up). Cumulative frequency of normal glucose tolerance in individuals with low risk (left anel, log-rank test p=0.03) and high risk (right panel, log-rank test 0.008). The inserts represent parametric survival models using fits of interval censored data. p=0.01 for the conventional vs. control intervention in the low-risk group (left panel) and p=0.003 for the intensive vs conventional intervention in the high risk group (right panel).

### Secondary outcomes (BMI, insulin sensitivity and secretion, liver fat content, cardiometabolic risk)

BMI decreased during LI (all p<0.0001), but not in controls (p=0.4). Liver fat content, cardiometabolic risk (Framingham score), insulin sensitivity and insulin secretion relative to insulin sensitivity only improved in high-risk subjects.

In high-risk subjects, liver fat, cardiometabolic risk, insulin sensitivity and BMI improved significantly more in the HR-INT group compared to the HR-CONV group after 1 year of LI, (see Table 2 and Figure 2 and 4, all p≤0.01). The change in insulin secretion was similar in the HR-CONV and HR-INT groups (p=0.8).

**Figure 4.**
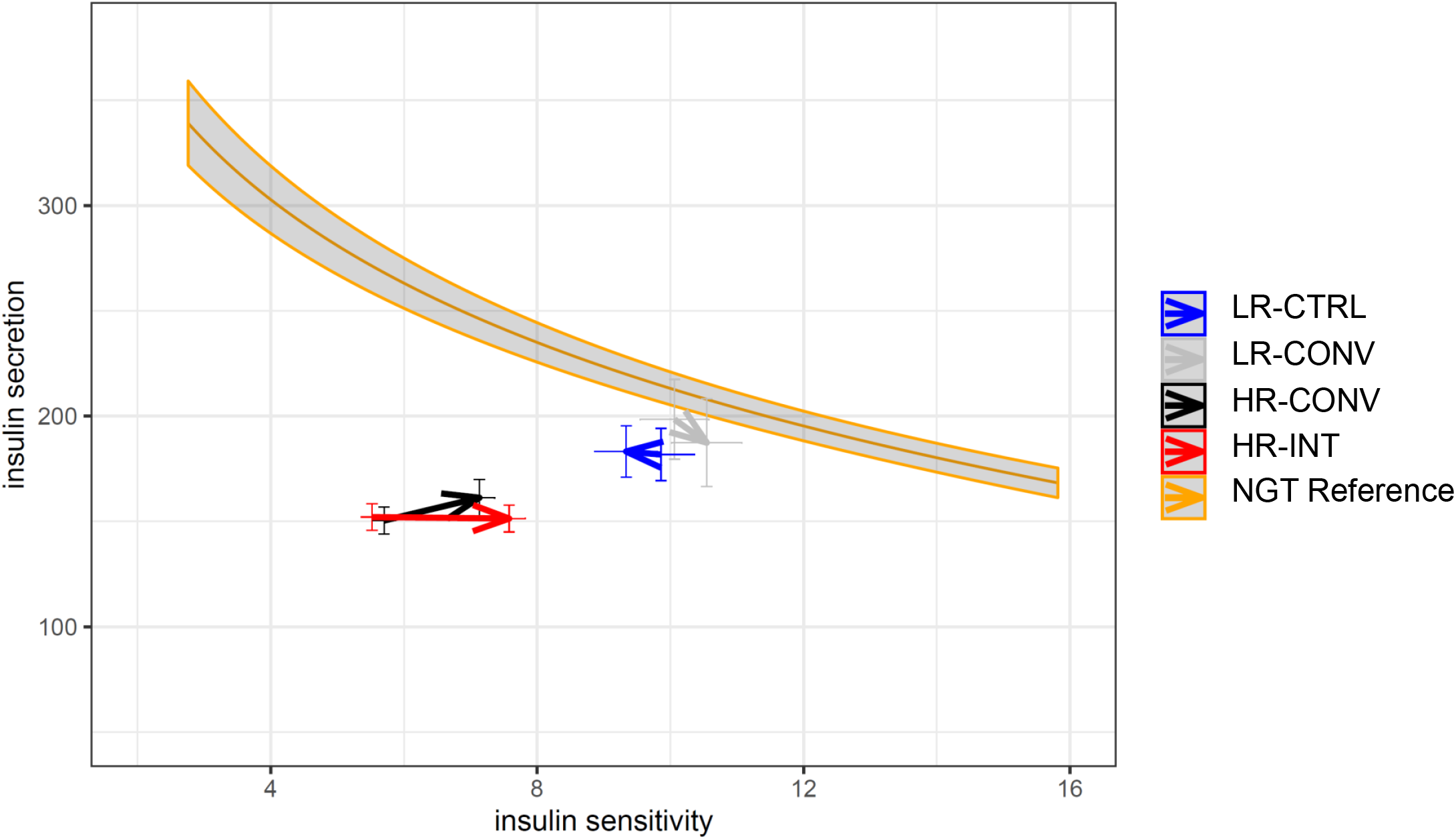
Change in insulin secretion and sensitivity during the study in the different study arms. Insulinogenic index (arbitrary units) as marker for insulin secretion is shown as hyperbolic function of insulin sensitivity (insulin sensitivity index, arbitrary units, unadjusted values). Subjects with normal glucose tolerance (NGT, age 18-50 years) from the German TUEF study (40) were used to compute the hyperbolic function (n=1421). The arrows represent baseline values (origin) and values after 12 months of intervention (end) in the study arms. 95 % CI are shown, blue: LR-CTRL, grey: LR-CONV, black: HR-CONV, red: HR-INT.

### Adherence

The aggregate percentage of completed lifestyle goals was higher in the LR-CONV than in the HR-CONV (45±3% vs 38±1%, p=0.03, Appendix Figure 3). In high-risk subjects, the aggregate percentage of completed lifestyle goals was similar in the HR-CONV (38±1%) compared to the HR-INT group (41±1%, p=0.5). When investigating the specific goals within the high-risk groups, more individuals reached exercise goals in the HR-CONV group, the weight goals were achieved by more individuals in the HR-INT group (both p<0.001, chi-squared test).

The main study outcome 2hPG was associated with the aggregate percentage of completed lifestyle goals (beta 0.15±0.04, p=0.001). In a multivariable model including specific lifestyle goals, only achievement of weight reduction (beta 0.38±0.06, p=0.04) and exercise goals (beta 0.13±0.06, p<0.001) were independently associated with glycemic improvement. In addition, the number of completed visits during the study was also positively associated with the improvement of glycemic improvement (beta 0.04±0.01, p=0.006).

### Safety and Adverse events

There were 0.88 adverse events per patient year. After adjusting for the number of visits and for centers, the frequency of adverse events was not different between LR-CONV and LR-CTRL groups, HR-INT and HR-CONV group as well as LR-CONV and HR-CONV group (all p>0.5, Poisson regression).

## Discussion

In the present multicenter, risk-stratified, randomized, controlled lifestyle intervention trial, our primary aim was to test whether individuals with prediabetes and a high-risk phenotype with impaired insulin secretion and/or insulin resistant fatty liver benefit from an intensification of LI. The PLIS study showed that in this population at high risk for diabetes, intensification of LI by increasing counselling frequency and weekly physical exercise indeed yielded a superior improvement of glucose metabolism after one year of LI. In addition, these participants undergoing intensive LI had by 62% higher transition probability into an improved glycemic category and had a higher cumulative frequency of regression to normal glucose tolerance during 3 years of follow up. They were also more probable to reduce secondary outcomes such as liver fat content and cardiometabolic risk.

This high-risk phenotype is defined by pathophysiological features of type 2 diabetes and has been described previously (12,16,17). The determinants of this phenotype, impaired insulin secretion and insulin resistance, are the main pathomechanisms for the development of type 2 diabetes (27-31). Notably, in our study insulin sensitivity improved only in individuals with this high-risk phenotype (see Figure 4).

Our data indicate that conventional lifestyle interventions, as were applied in the Diabetes Prevention Study (DPS) (1) and DPP (2), can be successfully intensified. This argues for a dose-effect relationship in LI. By applying the intensified intervention, beneficial effects on body fat mass, waist-to-hip ratio, insulin sensitivity, and liver fat content were more pronounced. In contrast, intensified LI did not improve insulin secretion capacity compared to conventional LI. Therefore, the superior effect of intensified LI on post-challenge glucose seems mainly due to reduced liver fat content and improved insulin sensitivity. The changes of liver fat content and insulin sensitivity were significantly associated with improvement of glucose tolerance, independent of change in body weight in the high-risk population (β=0.045, p=0.02 and β=-0.12, p<0.0001, respectively). The importance of improved insulin sensitivity in successful LI is consistent with findings from the DPP and DPS trials (32,33), whereas the data about the role of liver fat reduction is new.

The intensified and conventional intervention in PLIS differed in regard of exercise volume and the amount of counselling sessions. Of note, the number of completed visits and the accomplishment of the weight reduction goal were significantly associated with the reduction of 2hPG during one year of intervention in all treatment groups. This suggests that the amount of counselling and either more motivation or more guidance from lifestyle advisors underlies the higher efficacy of the intensive treatment group. Qualified lifestyle counsellors and an adequate counselling frequency should be key factors in LI planning. One additional important aspect are the perceptions and quality of life of participants taking part in the different lifestyle interventions. Quality of life during long term follow up and the feasibility of such lifestyle intervention in a real-world situation is being analysed in a separate project.

One accomplishment of the PLIS study is that we additionally tested the effect of conventional LI in the group with low-risk for LI non-response by comparing conventional LI with a group who did not receive any LI. No difference was found for the primary endpoint 2hPG between those groups. Although smaller sample size in the low-risk stratum might have precluded statistically significant differences, post-hoc sensitivity analyses with a bootstrap approach also suggest an intervention effect size that is approximately half of that in the high-risk stratum.

Several studies have shown that translating the promising results of controlled lifestyle interventions into a real-world scenario is hardly possible (34,35). Risk stratification during screening and subsequent allocation of resources to individuals who are at marked risk may improve outcomes and cost-effectiveness. For example, in individuals with type 2 diabetes, no advantage of a LI on cardiovascular disease mortality and morbidity was shown in the Look AHEAD trial (36). However, a post-hoc analysis has recently identified a subgroup that benefited from the LI. Individuals with well controlled diabetes (low risk) and poor self-reported general health did not benefit from the intervention (37). A screen and treat policy for the prevention of type 2 diabetes will be effective when it is possible to prospectively identify individuals at high-risk while excluding those at low risk (38). The current study provides a proof of concept for this approach.

Importantly, the beneficial effects of intensified LI reach beyond glucose control. The present study is the largest multicenter randomized LI trial measuring liver fat content with a highly reliable technique of magnetic resonance spectroscopy. Hepatic steatosis is present in 25% of the adult population in the United States, and is associated with diabetes, cardiovascular disease, steatohepatitis and liver cancer (31,39). In high-risk individuals, we achieved a relative liver fat reduction of 37% with intensified intervention, whereas conventional intervention only resulted in a relative reduction of 24%. Individuals who participated in the intensified intervention group achieved reduced liver fat content of 6.6±0.5% compared with 8.3±0.5% in those undergoing conventional intervention. This means that liver fat content was close to the normal threshold of 5.6% after the intensified intervention which therefore implies a clinically relevant effect and should be a target for future approaches to diabetes prevention.

Furthermore, the cardiovascular risk diminished in the participants of the high-risk stratum with a near doubling of risk reduction after intensified intervention, compared to conventional LI (see Table 2 and Figure 2).

Limitations of our study include the relative short LI duration of 12 months and a non-completer rate of 18% after one year. The latter is, however, well in the range of other LI studies with rates between 5% and 28% (5). A potential limitation is the heterogeneity of lifestyle counselling throughout different study centers which could have been reduced by more frequent meetings and interactions between study sites. Furthermore, the design of the present study did not include an intensified intervention in the low-risk group. Therefore, it may be possible that the level of physical activity was not sufficient to improve outcome in this group. In addition, there was no control intervention in the high-risk group. Moreover, the high risk and low risk group were unbalanced with more individuals stratified to the high-risk group (78%). Thus, one of the predefined questions “is lifestyle intervention effective in low-risk individuals with prediabetes” cannot be answered with high confidence in the current study.

To our knowledge, this is the first multicenter study that prospectively tested different intensities of lifestyle intervention in a risk-stratified manner. PLIS confirms the existence of a high-risk phenotype for non-response to LI in individuals with prediabetes. This non-response can be partially compensated with intensified LI such that a higher percentage of high-risk individuals improve glucose metabolism, decrease liver fat content and cardiovascular risk. Finally, conventional lifestyle intervention with the aim of improving glucose tolerance in individuals with prediabetes and low risk subphenotype might also be important but cannot be conclusively shown in our data. Generally, healthy lifestyle should certainly be promoted for other health benefits in all risk subphenotypes. Nonetheless, screen and treat approaches in the prevention of type 2 diabetes should include risk stratification and individualized interventions.

## Data Availability

Due to ethical regulations, we cannot share individual participant data. Mean values and confidence intervals of all analyzed patient-level data are available and maybe shared upon reasonable request. Study protocol, statistical analysis plan and analytic code used to generate results are available upon reasonable request. A request can be made to the corresponding author or officially to the German Center of Diabetes Research
For further information see data sharing statement included in the Supplementary Material.

## Contributors

AF, NS, and HUH conceived the study. AF, RW and ML analysed the data. AF and RW wrote the manuscript. AF, RW, MH, KK, PPN, AFP, MS, AB, HH, JS, AL, KW, SB, MR, NS, HUH contributed to interpretation of the data and edited the MS. All other authors contributed to data acquisition and approved the final version of the manuscript. AF is guarantor and attests all listed authors meet authorship criteria and no others meeting the criteria have been omitted. The lead author (AF) affirms that the manuscript is an honest, accurate, and transparent account of the study being reported and that no important aspects of the study have been omitted.

## Acknowledgments

We deeply thank all the study participants for their cooperation with this project. We gratefully acknowledge the excellent assistance and dedication of the study nurses, dietitians and lifestyle advisors in all the participating study sites. The study was supported by the DZD-German Center for Diabetes Research. The DZD is funded by the German Federal Ministry for Education and Research and the states where its partner institutions are located (01GI0925).

We acknowledge the authors of the following packages for the free statistical software R which were used in the data analysis: tidyverse, Hmisc, lsmeans, openxlsx, htmlTable, stringdist, mice, icenReg.

