## Supporting Information for "Risk-stratified lifestyle intervention to prevent type 2 diabetes"

This appendix has been provided by the authors to give readers additional information about their work.

Results of the randomized controlled Prediabetes Lifestyle Intervention Study (PLIS)

**Appendix Figure 1**

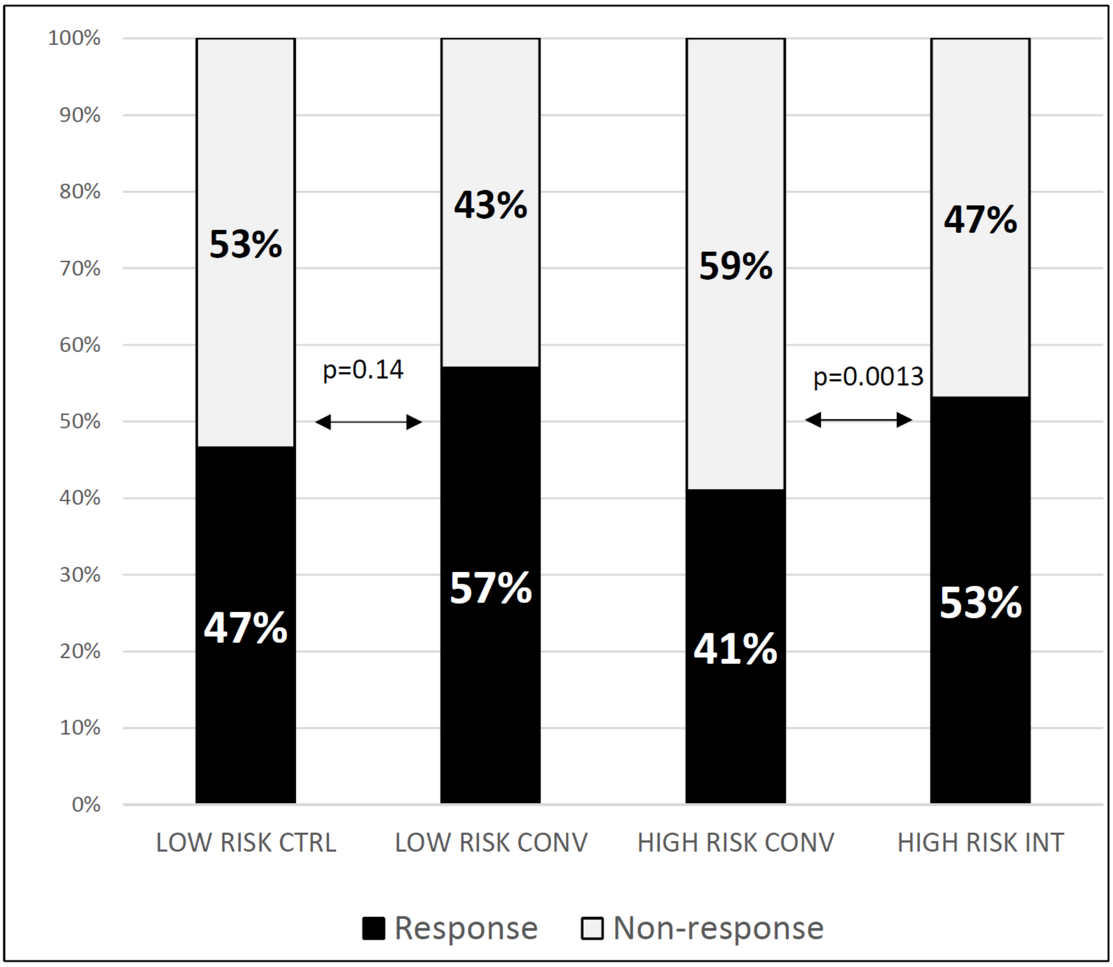

**Response and non-response to lifestyle intervention in the different intervention groups.**
CTRL = control, CONV = conventional lifestyle intervention, INT = intensified lifestyle intervention.

Response is defined as improvement of glucose regulation.

Specifically, participants with impaired fasting glucose (IFG) at baseline who have normal glucose tolerance (NGT) after intervention, participants with impaired glucose tolerance (IGT) at baseline who have NGT or IFG after intervention and participants with combined IFG+IGT at baseline who have NGT, isolated IFG or isolated IGT after intervention.

**Appendix Figure 2**

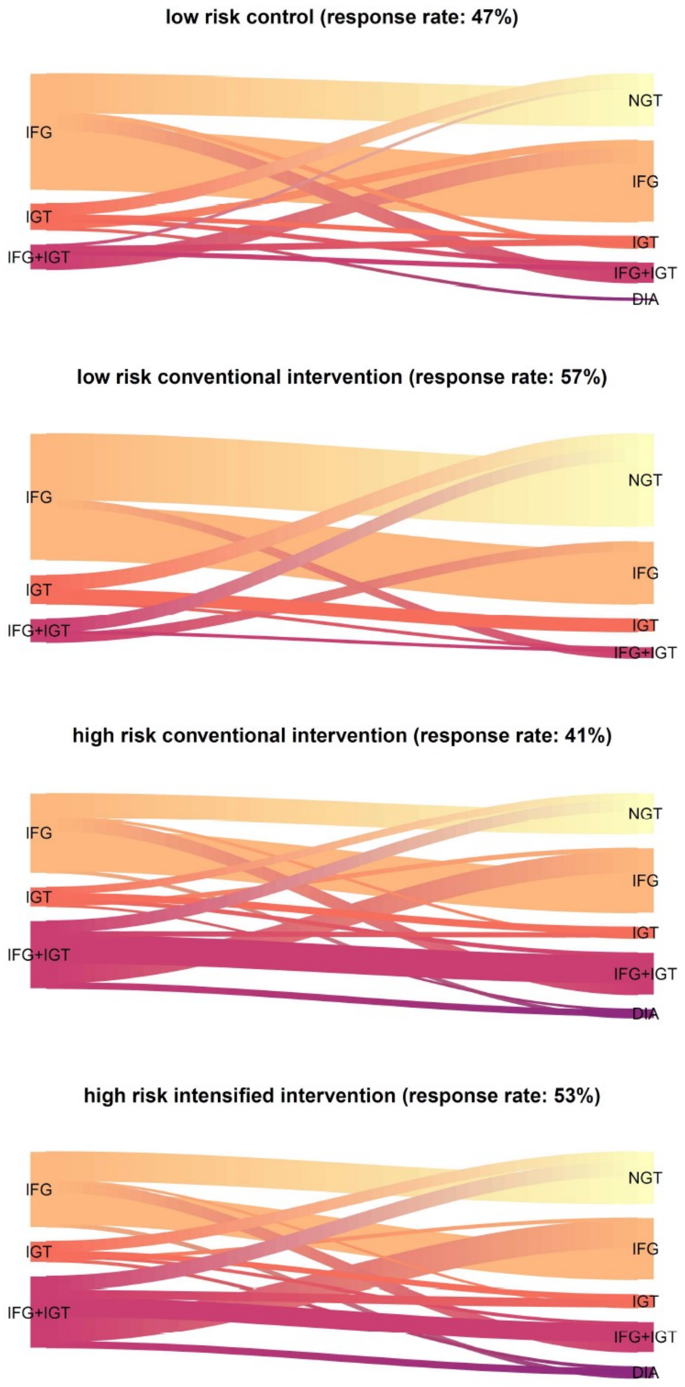

**Change in glucose tolerance categories from baseline to 12 months of LI and number of responders in the different risk and intervention groups.**

Response is defined as improvement of glucose regulation.

Specifically, participants with impaired fasting glucose (IFG) at baseline who have normal glucose tolerance (NGT) after intervention, participants with impaired glucose tolerance (IGT) at baseline who have NGT or IFG after intervention and participants with combined IFG+IGT at baseline who have NGT, isolated IFG or isolated IGT after intervention.

**Appendix Figure 3**

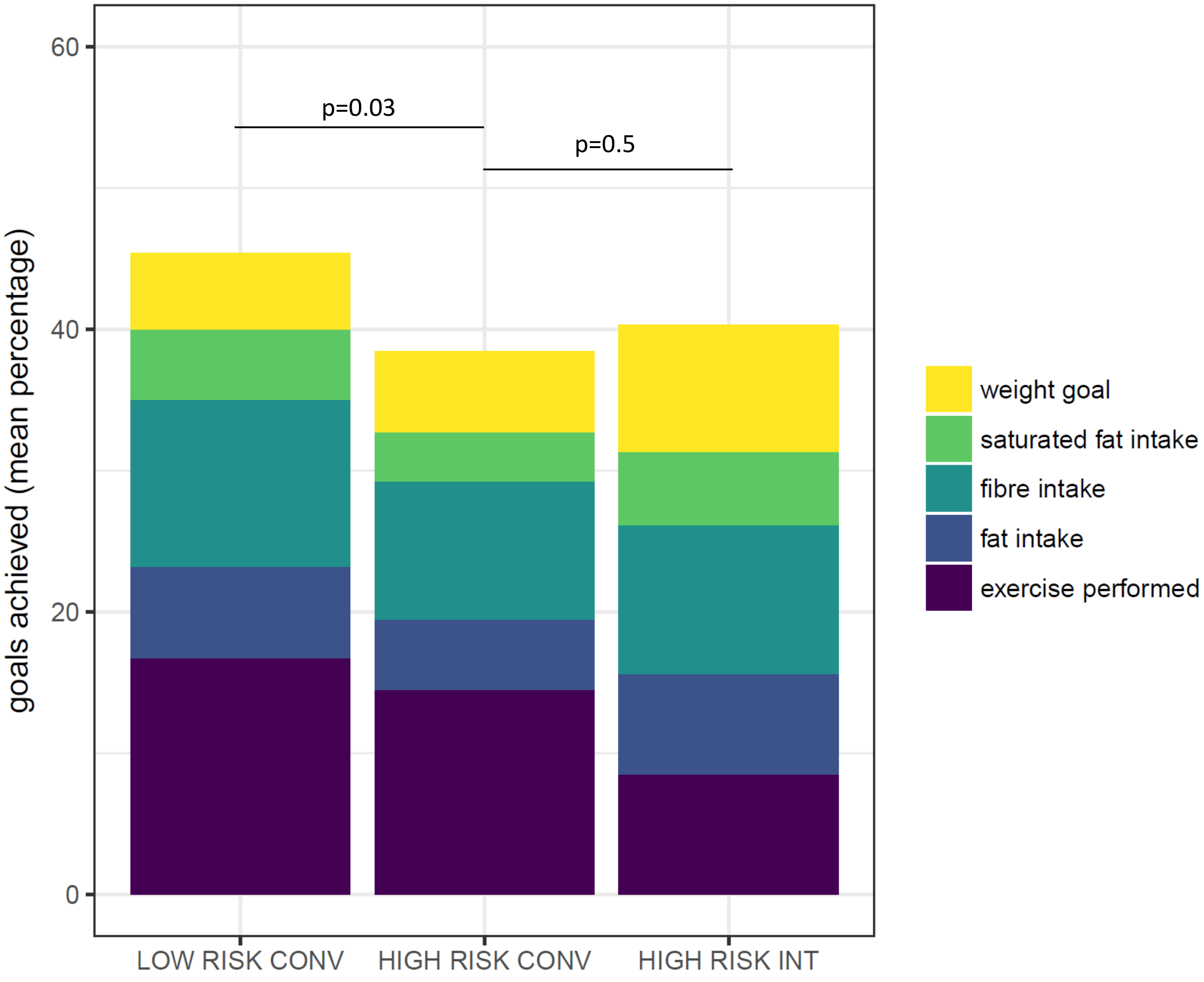

**Aggregate percentage of completed lifestyle goals during the study.**

To scale the aggregate percentage of all lifestyle goals to a maximum of 100%, the number of completed individual goals are downscaled by a factor of 5 (number of individual goals). The aggregate percentage of all completed lifestyle goals was higher in the LR-CONV compared to the HR-CONV (p=0.03, Wilcoxon-test) and not different between HR-CONV and HR-INT (p=0.5).

**Appendix Table 1**
**Participation at different academic diabetes centers where the study was performed.**

From each study center, about 2 fold of actually included individuals with prediabetes were potentially eligible for the study. For all study centers, a total of n=2561 were potentially eligible.

| | Number of participants receiving allocated intervention | Number of participants completing follow up |
|---|---|---|
| University Hospital of Tübingen | 351 | 304 |
| Universiy Hospital of Dresden | 213 | 178 |
| German Institute of Human Nutrition, Potsdam | 175 | 137 |
| University Hospital of Heidelberg | 104 | 80 |
| University of Düsseldorf | 59 | 48 |
| Technical University of Munich | 106 | 82 |
| Ludwig Maximilian University Munich | 49 | 40 |
| University Hospital of Leipzig | 48 | 39 |

**Appendix Table 2**
Exclusion criteria.

| Criterion | Specific definition |
|---|---|
| Pregnancy | |
| Lactation | |
| Symptomatic coronary artery disease | |
| Active malignant disease | unintended weight loss > 10% over the last 6 months |
| Elevated liver transaminases | 3 times above the upper limit of normal level |
| Chronic kidney disease | estimated glomerular filtration ratio < 50 $ml/min/1.73m^2$ |
| Systemic infection | |
| Glucocorticoid use | |
| Severe mental illness | |

**Appendix Table 3**
**Full-set analysis on all 1105 participants**
using multivariable imputation performed on a wide-dataset encompassing basic variables (sex, age, BMI, waist circumference, education, study center), and glycemic variables (glucose during OGTT, AUC glucose, HbA1c), variables on insulin secretion and sensitivity (ISI and IGI), disposition index as well as liver fat content at the visits at 6 months and 12 months. Missing data were imputed for all visits. The imputation was performed using the MICE package in R using default settings (predictive mean matching as default algorithm, 5 iterations) and passive imputation for derived variables (disposition indexes).

| | **LOW RISK** | | | **HIGH RISK** | | |
|---|---|---|---|---|---|---|
| | **LR-CTRL** | **LR-CONV** | **p-value** | **HR-CONV** | **HR-INT** | **p-value** |
| body mass index (kg $m^{-2}$) | 0 [-0.2;0.2] | -0.6 [-0.9;-0.4] | <0.0001 | -1.1 [-1.3;-1] | -1.6 [-1.8;-1.4] | <0.0001 |
| fasting glucose (mmol $l^{-1}$) | -0.13 [-0.21;-0.05] | -0.29 [-0.37;-0.21] | 0.004 | -0.2 [-0.25;-0.15] | -0.29 [-0.34;-0.24] | 0.02 |
| post-challenge glucose (mmol $l^{-1}$) | -0.49 [-0.76;-0.21] | -0.45 [-0.73;-0.18] | 0.9 | -0.65 [-0.82;-0.48] | -0.94 [-1.11;-0.77] | 0.02 |
| glucose AUC (mmol min $l^{-1}$) | -8 [-32;17] | -14 [-38;11] | 0.7 | -76 [-92;-61] | -105 [-120;-89] | 0.01 |
| glycated hemoglobin (%) | 0 [0;0] | 0 [-0.1;0] | 0.2 | -0.1 [-0.1;-0.1] | -0.1 [-0.2;-0.1] | 0.02 |
| Insulin sensitivity index | -1.2 [-1.8;-0.6] | -0.2 [-0.9;0.4] | 0.04 | 1.2 [0.9;1.5] | 1.9 [1.6;2.2] | 0.004 |
| Disposition index | -236 [-434;-37] | -247 [-446;-48] | 0.9 | 127 [51;203] | 168 [92;244] | 0.5 |
| Hepatic triglyceride content (%) | 0.1 [-0.3;0.5] | -0.1 [-0.5;0.3] | 0.4 | -3.1 [-3.5;-2.6] | -4.5 [-4.9;-4] | <0.0001 |

**Appendix Table 4:**
Comparison of baseline parameters (mean±SD), low-risk group control versus conventional intervention and high risk group conventional versus intensive intervention.

| | LR-CTRL | LR-CONV | HR-CONV | HR-INT |
|---|---|---|---|---|
| sex female/male (%) | 63/38 (62/38) | 61/39 (61/39) | 186/165 (53/47) | 209/147 59/41) |
| age (years) | 57±12 | 58±11 | 59±10 | 59±10 |
| weight (kg) | 80.1±16.1 | 81.3±16.3 | 92±19.7 | 92.4±19.2 |
| body mass index (kg $m^{-2}$) | 27.9±5.1 | 28.3±5.3 | 31.5±5.9 | 31.9±5.7 |
| waist circumference (cm) | 93±12 | 94±13 | 105±14 | 105±14 |
| waist-to-hip ratio | 0.89±0.09 | 0.89±0.08 | 0.94±0.09 | 0.94±0.08 |
| systolic blood pressure (mmHg) | 135±16 | 135±17 | 140±16 | 139±17 |
| diastolic blood pressure (mmHg) | 84±10 | 85±12 | 86±10 | 86±11 |
| fasting glucose (mmol $l^{-1}$) | 5.7±0.5 | 5.7±0.4 | 5.9±0.5 | 6.0±0.5 |
| post-challenge glucose (mmol $l^{-1}$) | 6.9±1.4 | 6.7±1.5 | 7.8±1.7 | 7.8±1.7 |
| glucose AUC (mmol min $l^{-1}$) | 935±112 | 933±130 | 1131±160 | 1131±161 |
| glycated hemoglobin (%) | 5.7±0.3 | 5.6±0.3 | 5.8±0.3 | 5.8±0.4 |
| glycated hemoglobin (mmol $mol^{-1}$) | 38.4±3.6 | 37.8±3.6 | 40.1±3.5 | 39.4±4.1 |
| triglycerides (mmol $l^{-1}$) | 1.26±0.96 | 1.24±0.72 | 1.64±0.95 | 1.63±0.98 |
| cholesterol (mmol $l^{-1}$) | 5.34±0.86 | 5.22±0.87 | 5.46±1.09 | 5.43±1.02 |
| LDL cholesterol (mmol $l^{-1}$) | 3.23±0.83 | 3.12±0.79 | 3.38±0.95 | 3.3±0.87 |
| HDL cholesterol (mmol $l^{-1}$) | 1.47±0.4 | 1.56±0.71 | 1.37±0.36 | 1.39±0.41 |
| Liver fat content (%) | 2.85±2.74 | 2.86±3.1 | 10.72±8.76 | 10.18±7.57 |
| Insulin sensitivity index (AU) | 9.86±5.05 | 10.06±5.16 | 5.70±3.12 | 5.52±3.01 |
| Insulin secretion (Disposition index) (AU) | 1440±994 | 1627±1352 | 654±424 | 688±506 |
| Hypertension no/yes (%) | 56/39 (59/41) | 63/33 (66/34) | 155/185 (46/54) | 152/181 (46/54) |

| | | | | |
|---|---|---|---|---|
| Hyperlipidemia no/yes (%) | 55/38 (59/41) | 58/34 (63/37) | 184/145 (56/44) | 171/145 (54/46) |
| History of myocardial infarction no/yes (%) | 91/3 (97/3) | 97/1 (99/1) | 325/7 (98/2) | 314/10 (97/3) |
| History of stroke no/yes (%) | 90/4 (96/4) | 95/2 (98/2) | 325/4 (99/1) | 311/13 (96/4) |
| peripheral artery disease no/yes (%) | 85/6 (93/7) | 88/7 (93/7) | 288/41 (88/12) | 284/39 (88/12) |
| medication: angiotensine convertase inhibitors no/yes (%) | 89/12 (88/12) | 91/9 (91/9) | 290/61 (83/17) | 303/53 (85/15) |
| medication: angiotensine receptor blockers no/yes (%) | 84/17 (83/17) | 84/16 (84/16) | 276/75 (79/21) | 260/96 (73/27) |
| medication: thiazide diuretics no/yes (%) | 93/8 (92/8) | 92/8 (92/8) | 306/45 (87/13) | 301/55 (85/15) |
| medication: other diuretics no/yes (%) | 98/3 (97/3) | 98/2 (98/2) | 335/16 (95/7) | 341/15 (96/4) |
| medication: beta blockers no/yes (%) | 88/13 (87/13) | 86/14 (86/14) | 272/79 (77/23) | 273/83 (77/23) |
| medication: statins no/yes (%) | 89/12 (88/12) | 86/14 (86/14) | 287/64 (82/18) | 294/62 (83/17) |
| current smoking no/yes (%) | 93/3 (97/3) | 91/7 (93/7) | 324/22 (94/6) | 321/22 (94/6) |
| alcohol consumption n (%) | | | | |
| 1_none | 20 (21) | 11 (11) | 29 (8) | 42 (12) |
| 2_rarely | 30 (31) | 43 (44) | 157 (45) | 147 (43) |
| 3_week-ends | 9 (9) | 5 (5) | 33 (10) | 24 (7) |
| 4_weekely_2-3 | 33 (34) | 27 (27) | 82 (24) | 87 (26) |
| 5_daily | 5 (5) | 13 (13) | 44 (13) | 40 (12) |
| highest education n (%) | | | | |
| 1_none | 1 (1) | 4 (4) | 8 (2) | 11 (3) |
| 2_post_secondary | 46 (47) | 53 (54) | 160 (48) | 154 (45) |
| 3_bachelor_or_equivalent | 18 (18) | 15 (15) | 85 (25) | 89 (26) |
| 4_master_or_equivalent | 34 (34) | 26 (27) | 84 (25) | 88 (26) |

**Appendix Table 5**

Comparison of baseline variables (mean±SD) non-completers versus completers.

| | Non-completer (n=197) | Completer (n=908) | p-value |
|---|---|---|---|
| Riskgroup n (%) | | | 1 |
| 1_low risk | 44 (22) | 201 (22) | |
| 2_high risk | 153 (78) | 707 (78) | |
| Intervention group n (%) | | | 0.91 |
| control | 22 (11) | 101 (11) | |
| conventional | 101 (51) | 451 (50) | |
| intensive | 74 (38) | 356 (39) | |
| sex female/male (%) | 132/65 (67/33) | 519/389 (57/43) | 0.014 |
| age (years) | 54±12 | 59±10 | <0.0001 |
| weight (kg) | 93.1±21.1 | 89.6±19.4 | 0.036 |
| body mass index (kg $m^{-2}$) | 32.2±6.5 | 30.9±5.9 | 0.0091 |
| waist circumference (cm) | 104±16 | 102±14 | 0.19 |
| waist-to-hip ratio | 0.92±0.09 | 0.93±0.09 | 0.19 |
| systolic blood pressure (mmHg) | 136±16 | 139±17 | 0.048 |
| diastolic blood pressure (mmHg) | 86±10 | 86±11 | 0.98 |
| fasting glucose (mmol $l^{-1}$) | 6.0±0.6 | 5.9±0.5 | 0.089 |
| post-challenge glucose (mmol $l^{-1}$) | 7.5±1.9 | 7.6±1.7 | 0.7 |
| glucose AUC (mmol min $l^{-1}$) | 1084±191 | 1088±173 | 0.79 |
| glycated hemoglobin (mmol $mol^{-1}$) | 39.3±4.4 | 39.4±3.8 | 0.72 |
| glycated hemoglobin (%) | 5.7±0.4 | 5.8±0.3 | 0.72 |
| triglycerides (mmol $l^{-1}$) | 1.53±0.86 | 1.55±0.95 | 0.83 |
| cholesterol (mmol $l^{-1}$) | 5.33±0.98 | 5.41±1.01 | 0.31 |
| LDL cholesterol (mmol $l^{-1}$) | 3.26±0.88 | 3.3±0.89 | 0.52 |
| HDL cholesterol (mmol $l^{-1}$) | 1.39±0.38 | 1.41±0.44 | 0.59 |
| Hepatic triglyceride content (%) | 9.12±8.21 | 8.75±8 | 0.62 |
| Insulin sensitivity index | 6.38±3.68 | 6.58±4.04 | 0.52 |
| Insulin secretion (Disposition index) (AU) | 882.±660 | 863±781 | 0.73 |
| Hypertension no/yes (%) | 95/84 (53/47) | 426/438 (49/51) | 0.4 |
| hyperlipidemia no/yes (%) | 108/64 (63/37) | 468/362 (56/44) | 0.14 |
| myocardial infarction no/yes (%) | 172/4 (98/2) | 827/21 (98/2) | 1 |
| stroke no/yes (%) | 176/2 (99/1) | 821/23 (97/3) | 0.32 |
| peripheral artery disease no/yes (%) | 154/21 (88/12) | 745/93 (89/11) | 0.83 |
| medication: angiotensine convertase inhibitors no/yes (%) | 171/26 (87/13) | 773/135 (85/15) | 0.62 |
| medication: angiotensine receptor blockers no/yes (%) | 160/37(81/19) | 704/204 (78/22) | 0.3 |
| medication: thiazide diuretics no/yes (%) | 176/21 (89/11) | 792/116 (87/13) | 0.49 |
| medication: other diuretics no/yes (%) | 194/3 (98/2) | 872/36 (96/4) | 0.14 |
| medication: beta blockers no/yes (%) | 156/41 (79/21) | 719/189 (79/21) | 1 |

| | | | |
|---|---|---|---|
| medication: statins no/yes (%) | 173/24 888/12) | 756/152 (83/17) | 0.14 |
| current smoking no/yes (%) | 154/26 (86/14) | 829/54 (94/6) | 0.00021 |
| alcohol use | | | 0.025 |
| 1_none | 31 (17) | 102 (12) | |
| 2_rarely | 89 (50) | 377 (42) | |
| 3_week-ends | 13 (7) | 71 (8) | |
| 4_weekely_2-3 | 31 (17) | 229 (26) | |
| 5_daily | 16 (9) | 102 (12) | |
| highest education | | | 0.62 |
| 1_none | 6 (3) | 24 (3) | |
| 2_post_secondary | 95 (52) | 413 (47) | |
| 3_bachelor_or_equivalent | 38 (21) | 207 (24) | |
| 4_master_or_equivalent | 44 (24) | 232 (26) | |

**Appendix Table 6**

Effect of conventional LI in low risk individuals versus high risk individuals.

*ANCOVA, adjusted for baseline and center.

| | **Between-group difference of conventional intervention * (for HR-CONV, reference: LR-CONV beta coefficient (±SE))** | **p-value** |
|---|---|---|
| age (years) | -0 (±0) | 0.6 |
| weight (kg) | 0.4 (±0.6) | 0.5 |
| body mass index (kg $m^{-2}$) | 0.1 (±0.2) | 0.5 |
| waist circumference (cm) | -1 (±1) | 0.6 |
| waist-to-hip ratio | -0.02 (±0.01) | 0.008 |
| lean mass percent | -5.5 (±2.3) | 0.02 |
| fat mass percent | 0.9 (±0.5) | 0.07 |
| habitual physical activity score | 0.1 (±0.1) | 0.5 |
| systolic blood pressure (mmHg) | -3 (±2) | 0.09 |
| diastolic blood pressure (mmHg) | -1 (±1) | 0.3 |
| heart rate (1/min) | -0 (±1) | 0.7 |
| fasting glucose (mmol $l^{-1}$) | -0.16 (±0.06) | 0.006 |
| post-challenge glucose (mmol $l^{-1}$) | -0.55 (±0.20) | 0.008 |
| glucose AUC (mmol min $l^{-1}$) | -33 (±20) | 0.1 |
| glycated hemoglobin (mmol $mol^{-1}$) | -0.0 (±0.4) | 1 |
| glycated hemoglobin (%) | -0.0 (±0.0) | 1 |
| triglycerides (mmol $l^{-1}$) | 0.11 (±0.07) | 0.1 |
| cholesterol (mmol $l^{-1}$) | 0.12 (±0.08) | 0.1 |
| LDL cholesterol (mmol $l^{-1}$) | 0.09 (±0.07) | 0.2 |
| HDL cholesterol (mmol $l^{-1}$) | 0.04 (±0.03) | 0.2 |
| C-reactive protein (mg $dl^{-1}$) | 0.1 (±0.8) | 0.9 |
| Aspartate aminotransferase (Units $l^{-1}$) | 0.1 (±1.4) | 0.9 |
| Alanin aminotransferase (Units $l^{-1}$) | 0.3 (±1.5) | 0.8 |
| Gamma glutamyltransferase (Units $l^{-1}$) | 2.4 (±2.1) | 0.3 |
| Insulin sensitivity index | -0.1 (±0.5) | 0.9 |
| Insulin secretion (Disposition index) (AU) | 226 (±134) | 0.09 |
| Liver fat content (%) | 0.1 (±0.6) | 0.9 |
| Framingham 10-year-CV-risk (%) | -0.4 (±1.0) | 0.7 |

**Participating Investigators**

University Hospital Tübingen:
Andreas Fritsche, Robert Wagner, Martin Heni, Kostantinos Kantartzis, Jürgen Machann, Fritz Schick, Rainer Lehmann, Andreas Peter, Corinna Dannecker, Louise Fritsche, Vera Valenta, Norbert Stefan, Hans-Ulrich Häring

DZD Head Office:
Renate Schick

University Hospital Heidelberg:
Peter Paul Nawroth, Stefan Kopf

German Institute of Human Nutrition:
Andreas FH Pfeiffer, Stefan Kabisch, Ulrike Dambeck, Annette Schürmann

University of Leipzig:
Michael Stumvoll, Matthias Blüher

Technical University Dresden:
Andreas L Birkenfeld, Peter Schwarz, Stefan Bornstein

Technical University Munich:
Hans Hauner, Julia Clavel

Ludwig-Maximilians University Munich:
Jochen Seißler, Andreas Lechner

Heinrich-Heine University Düsseldorf:
Karsten Müssig, Katharina Weber, Michael Roden

Helmhotz Center Munich:
Michael Laxy, Martin Hrabe de Angelis

**Data Sharing Statement**

Individual participant data cannot be shared publicly because of ethical regulations. Mean values and confidence intervals of all analyzed patient-level data are available for researchers who meet the criteria for access to confidential data. Data are available upon request to the corresponding author or officially to the German Center of Diabetes Research.

**Financial information**

Funding: The DZD is funded by the German Federal Ministry for Education and Research (01GI0925).

**Detailed explanation for why the trial was registered late in clinicaltrials.gov.**

A timely registration of the PLIS trial in a United States registry of clinical trials has been unfortunately been overlooked by us during the initiation of the study in 2012, and was completed only in 2013. We apologize for this unintentional delayed registration at "clinicaltrials.gov". However, the whole study protocol, also comprising all end-points, was authorized and registered by the ethical committees of all participating centers (n=8) prior to recruiting participants. The protocol had been publicly available in German language for all researchers within the German Center for Diabetes Research (DZD) network from the beginning of 2012, i.e. well before the first patient entered the study. We then initiated a translation of the German study protocol and organized a submission to "clinicaltrials.gov." which unfortunately has been performed with delay.
The principal investigator and the speakers of the German Center for Diabetes Research, as well as the institutional review boards can confirm that there were no changes in the study protocol from 2012 up until 2015. All subsequent (minor) changes were also documented at clinicaltrials.gov (2015 and 2017). Therefore, we are confident that the integrity of our data is not affected by the delayed registration at clinicaltrials.gov.
The precise date of trial registration and the date the first participant was enrolled are as follows: The first patient was enrolled on May 9, 2012. The trial was submitted on June 25, 2013 to clinical trials .gov
From May 9, 2012 to June 25, 2013, we enrolled 89 patients over all study centers, this is 9,8% of the final analyzed study population (n=908)
There are no discrepancies between the Institutional Review Board application and the clinical trials.gov registration.

CONSORT 2010 checklist of information to include when reporting a randomised trial*

| Section/Topic | Item No | Checklist item | Reported on page No |
|---|---|---|---|
| **Title and abstract** | | | |
| | 1a | Identification as a randomised trial in the title | Title page, p. 1 |
| | 1b | Structured summary of trial design, methods, results, and conclusions (for specific guidance see CONSORT for abstracts) | Abstract (Methods and Findings), p. 3 |
| **Introduction** | | | |
| Background and objectives | 2a | Scientific background and explanation of rationale | Abstract p. 3, Intro, p. 6/7 |
| | 2b | Specific objectives or hypotheses | Abstract p. 3, Intro, p. 7 |
| **Methods** | | | |
| Trial design | 3a | Description of trial design (such as parallel, factorial) including allocation ratio | Abstract p. 3, Methods: Study design, p.8 |
| | 3b | Important changes to methods after trial commencement (such as eligibility criteria), with reasons | n.a., no changes |
| Participants | 4a | Eligibility criteria for participants | Abstract p. 3 Methods, p. 8/9 (Participants) |
| | 4b | Settings and locations where the data were collected | Abstract (p. 3) Methods, p.8, Appendix table 1 |
| Interventions | 5 | The interventions for each group with sufficient details to allow replication, including how and when they were actually administered | Abstract p. 3, Methods, p. 9 (Intervention) |
| Outcomes | 6a | Completely defined pre-specified primary and secondary outcome measures, including how and when they were assessed | Abstract p. 3, Methods, p. 8, 10, 11 |
| | 6b | Any changes to trial outcomes after the trial commenced, with reasons | n.a. no changes |
| Sample size | 7a | How sample size was determined | Methods, p. 11 (Statistical analysis) |
| | 7b | When applicable, explanation of any interim analyses and stopping guidelines | n.a. no interim analyses |
| Randomisation: | | | |
| Sequence generation | 8a | Method used to generate the random allocation sequence | Methods, p. 8 (study design) |
| | 8b | Type of randomisation; details of any restriction (such as blocking and block size) | Methods, p. 8 (study design) |

| | | | |
|---|---|---|---|
| Allocation concealment mechanism | 9 | Mechanism used to implement the random allocation sequence (such as sequentially numbered containers), describing any steps taken to conceal the sequence until interventions were assigned | Methods, p.8 (study design) |
| Implementation | 10 | Who generated the random allocation sequence, who enrolled participants, and who assigned participants to interventions | Methods p. 8 (study design) |
| Blinding | 11a | If done, who was blinded after assignment to interventions (for example, participants, care providers, those assessing outcomes) and how | Methods, p.8 (study design) |
| | 11b | If relevant, description of the similarity of interventions | n.a. |
| Statistical methods | 12a | Statistical methods used to compare groups for primary and secondary outcomes | Methods, p.11 (Statistical analysis) |
| | 12b | Methods for additional analyses, such as subgroup analyses and adjusted analyses | Methods, p.11 |
| **Results** | | | |
| Participant flow (a diagram is strongly recommended) | 13a | For each group, the numbers of participants who were randomly assigned, received intended treatment, and were analysed for the primary outcome | Results, p. 12-14 |
| | 13b | For each group, losses and exclusions after randomisation, together with reasons | Results, p.12 and Fig. 1 |
| Recruitment | 14a | Dates defining the periods of recruitment and follow-up | Methods, p.8 (study design) |
| | 14b | Why the trial ended or was stopped | Methods, p. 8 (study design) |
| Baseline data | 15 | A table showing baseline demographic and clinical characteristics for each group | Table 1 |
| Numbers analysed | 16 | For each group, number of participants (denominator) included in each analysis and whether the analysis was by original assigned groups | Fig. 1 and Appendix Table 3 |
| Outcomes and estimation | 17a | For each primary and secondary outcome, results for each group, and the estimated effect size and its precision (such as 95% confidence interval) | Abstract p. 3, Results, p.12-14, Fig. 2-4 |
| | 17b | For binary outcomes, presentation of both absolute and relative effect sizes is recommended | Results, p.12-14, |
| Ancillary analyses | 18 | Results of any other analyses performed, including subgroup analyses and adjusted analyses, distinguishing pre-specified from exploratory | Results p. 13, (changes in glycemic category) |
| Harms | 19 | All important harms or unintended effects in each group (for specific guidance see CONSORT for harms) | Methods, p. 15 |
| **Discussion** | | | |
| Limitations | 20 | Trial limitations, addressing sources of potential bias, imprecision, and, if relevant, multiplicity of analyses | Discussion, p. 19 |
| Generalisability | 21 | Generalisability (external validity, applicability) of the trial findings | Discussion, p. 19 |
| Interpretation | 22 | Interpretation consistent with results, balancing benefits and harms, and considering other relevant evidence | Discussion, p. 18/19 |

| | | | |
|---|---|---|---|
| **Other information** | | | |
| Registration | 23 | Registration number and name of trial registry | Abstract, p. 4 |
| Protocol | 24 | Where the full trial protocol can be accessed, if available | Methods, p. 8 |
| Funding | 25 | Sources of funding and other support (such as supply of drugs), role of funders | Information pulled from article meta-data |

*We strongly recommend reading this statement in conjunction with the CONSORT 2010 Explanation and Elaboration for important clarifications on all the items. If relevant, we also recommend reading CONSORT extensions for cluster randomised trials, non-inferiority and equivalence trials, non-pharmacological treatments, herbal interventions, and pragmatic trials. Additional extensions are forthcoming: for those and for up to date references relevant to this checklist, see www.consort-statement.org.
